# Addressing the Challenges with Non-Communicable Diseases among People Living with HIV: A Mixed Methods Research on Burden, Barriers, and Future Directions in a Tertiary Health Care Institution in India

**DOI:** 10.1101/2025.07.01.25329897

**Authors:** Sandipta Chakraborty, Shantasil Pain, Pankaj Kumar Mandal, Bobby Paul, Dipankar Jana, Preeti Gurung

**Author notes:** **Name and address (and email) of the author correspondence:** Sandipta CHAKRABORTY, 187 Jonepur Road, Kanchrapara, North 24 PGS, West Bengal, Pin-743145 India.

## Abstract

**Background:** As the demographic of people living with HIV (PLHIV) ages, this population is increasingly experiencing a dual burden of HIV and non-communicable diseases (NCDs), alongside significant challenges in seeking NCD care. This study estimated the burden and risk factors of common NCDs including diabetes mellitus (DM), Hypertension (HTN), heart disease, stroke, cancer, and respiratory disease, among PLHIV, explored NCD-care seeking behavior and barriers, and feasibility of integrated HIV-NCD services.

**Methods:** Convergent parallel mixed-methods research with quantitative cross-sectional strand among 279 systematically selected adult PLHIV at an antiretroviral therapy (ART) center, captured NCD burden and healthcare seeking behavior. In-depth interviews (IDIs) of 10 PLHIV with NCDs and three key informant interviews (KIIs) with ART service providers, in qualitative strand explored the care seeking pattern, barriers and the potential of comprehensive HIV-NCD care.

**Results:** Almost 30% of PLHIV reported NCDs, predominantly DM (14.0%) and HTN (13.3%). Older age, physical inactivity, and obesity were significant risk factors. NCD care was preferred from the private health sector (59%), more in case of multimorbidity (66.7%). Qualitative exploration revealed that individual, interpersonal, and community-organizational level barriers hinder the NCD care among PLHIV. Public health care suffered from system fragmentation, while private care carries financial risk.

**Conclusions:** The aging PLHIV faces a substantial burden of NCD compounded by numerous barriers to seeking health care, like fragmented healthcare systems, financial constraints, physical inability etc. This complex issue warrants a comprehensive HIV-NCD service within public health systems. New, patient-centered models of integrated care warrant further investigation.

**What is already known on this topic:** With improved life expectancy, the aging people living with HIV (PLHIV) are at risk of suffering from NCDs and subsequently the chronic nature of both HIV and NCDs can disproportionately affect them. Both diseases need sustainable health care delivery. PLHIV sometimes faces additional barriers while seeking healthcare for NCDs. However, in Indian context, the magnitude of NCDs, provision of health care delivery and barriers to seek care among the PLHIV is not well evidenced.

**What this study adds:** The present study revealed the magnitude of common NCDs and known risk factors among a representative PLHIV population, attending an antiretroviral therapy (ART) center in India. Furthermore, this study added the healthcare seeking behavior of PLHIV for NCDs. Barriers to NCD care for PLHIV were explored through the perspective of both the PLHIV and healthcare providers. The study concluded with the need of comprehensive HIV-NCD care and discuss the avenues to implement the same in Indian context.

**How this study might affect research, practice, or policy:** At policy level the present public health system in India envisions a comprehensive chronic care model for PLHIV embedding screening and treatment for NCDs. However, implementation challenges exist. In practice, addressing fragmentation within the existing system could be an opportunity to improve NCD interventions for PLHIV. Further research into implementing new models of comprehensive HIV-NCD care within the available healthcare resources is warranted through the current findings.

## Introduction

Noncommunicable diseases (NCDs) are now the leading global contributors to morbidity, mortality, and out-of-pocket healthcare expenses. Four main categories of NCDs, cardiovascular diseases (CVDs; like heart attacks and strokes), cancer, chronic respiratory diseases (like chronic obstructive pulmonary disease and asthma), and diabetes mellitus (DM) significantly impact global health outcomes. Hypertension (HTN), a major risk factor for CVDs, also contributes substantially to NCD-related illness and death [1, 2].

Individuals living with human immunodeficiency virus (HIV) infection and acquired immune deficiency syndrome (AIDS) are no exception to this. The chronicity of both HIV and NCDs can disproportionately impact people living with HIV (PLHIV) [3]. Advances in antiretroviral therapy (ART) and public health interventions resulted in improved life expectancy, aging and expansion in global PLHIV population [4]. PLHIV older than 50 years was estimated to exceed 20% by 2020, compared to 8% reported in 2000 [5]. Alike general population, older PLHIV are at higher risk of developing NCDs [6, 7]. Known risk factors for NCDs, including obesity, addiction, physical inactivity, and poor dietary habits, are highly prevalent among PLHIV [8]. Additionally, HIV infection may accelerate aging process and the early onset of NCDs [9].

Globally, studies indicated a broad range of NCD burden among PLHIV (14% to 48%), with HTN and DM being the most frequent [6, 10–12]. The co-occurrence of NCDs among PLHIV imposes a dual burden, necessitating lifelong care for both conditions. Unfortunately, PLHIV face numerous additional barriers when seeking healthcare for NCDs including physical disabilities, economic constraints, transportation challenges, and fragmented healthcare delivery systems. Furthermore, stigma and discrimination also impede effective delivery of NCD care to this population [13–15].

In India, despite low adult HIV prevalence (0.21%), the absolute number of PLHIV is substantial and features an increasingly aging demographic [16]. Given this large number of aging PLHIV and the overall high prevalence of NCDs, India is expected to have many individuals requiring lifelong care for both HIV and NCDs. The recent National Guidelines for HIV Care and Treatment in India from the National AIDS Control Organization (NACO), emphasize on a comprehensive care approach for the aging PLHIV population and provide specific recommendations for the prevention and management of NCDs [17]. However, there remains a dearth of evidence in India concerning the struggle with NCDs among PLHIV including the magnitude of dual burden, barriers to NCD care seeking and the nature of healthcare delivery offered for NCDs to the PLHIV. In response to this knowledge gap this study was designed to estimate the burden and identify risk factors for common NCDs, such as DM, HTN, heart disease, stroke, cancer, and respiratory disease, among PLHIV. Furthermore, the study also explored the patterns of NCD healthcare seeking behavior and identified potential barriers to care. Based on these findings, the feasibility of comprehensive care model for HIV-NCD is discussed.

## Methods

Convergent parallel mixed-methods research (MMR) design was employed. The primary, quantitative component (QUAN), with a cross-sectional approach recruited PLHIV attending a randomly selected ART center of a tertiary care hospital. Eligible participants were aged ≥18 years, and on ART for ≥ 6 months. Critically ill, bedridden, individuals were excluded.

The secondary, qualitative strand (Qual) engaged PLHIV from the same ART center, who had been living with at least one NCD for ≥ 6 months duration. Additionally, ART service providers were included as Key Informants (KIs).

This MMR approach was essential for capturing comprehensive insights from both PLHIV and healthcare providers regarding the NCD burden, service provision, healthcare-seeking behaviors, and barriers to care. Data were collected in sequence, analyzed simultaneously (QUAN_da_→←Qual_da_), triangulated and integrated [18].

Self-reported DM, HTN, heart diseases, stroke, cancer, and respiratory diseases, were collectively referred as NCDs. Multimorbidity was defined by presence of more than one NCDs. A minimum sample size of 282 was calculated with Cochran’s formula, accounting previously reported NCD burden among PLHIV (20.7%), alpha error (5%), absolute precision (10%), and anticipated nonresponse rate (10%) [12]. Participants were selected using systematic random sampling, assuming equivalence to simple random sampling due to the absence of a fixed order or periodicity among ART attendees. Sampling interval (50/6 ≈ 8) was calculated based on estimated daily average of 50 ART attendees and a feasible target of six interviews per day. Daily recruitment began by randomly selecting a patient between the first and eighth positions in the daily ART register, followed by approaching every subsequent eighth individual until six interviews were completed [19]. Written informed consent was obtained from all selected individuals. In case of ineligibility or refusal, the next ART attendee in the sequence was approached.

Consenting participants were interviewed using a pre-tested, structured questionnaire administered in local language capturing information on socio-demography, addictions (smoking, tobacco use, and alcohol consumption), physical activity, dietary habits, self-reported NCDs and healthcare-seeking behaviors [20, 21].

Baseline and recent CD4+ count and latest HIV viral load, were extracted from ART center medical records. Body Mass Index (BMI) was calculated from anthropometric measurements (height and weight) [20]. Data were entered using Epicollect 5, anonymized and coded during analysis.

Quantitative data were summarized using descriptive statistics; including number, percentage, mean, median, standard deviation (SD), interquartile range (IQR), and are presented in tables and figures. Logistic regression analysis was performed to identify factors associated with NCDs, using unadjusted and adjusted odds ratios (uOR and aOR) with 95% confidence intervals (95% CI). Quantitative analyses were performed with Stata 17.0

Qualitative data were generated through in-depth interviews (IDIs) with PLHIV suffering from NCDs and key informant interviews (KIIs) with ART service providers. PLHIV were purposively sampled ensuring maximum variation across genders, and presence of single versus multiple NCDs with recruitment continuing until saturation of information. A pre-designed and tested IDI guide was used to explore NCD associated healthcare-seeking behaviors and barriers. McLeroy’s socioecological model (SEM) was adapted to address barriers at individual, interpersonal, and community-organizational levels [22]. ART service providers, including medical doctors, nursing officers, and counsellors, were conveniently sampled and interviewed using a semi-structured KII guide, additionally focusing on the necessity and feasibility of comprehensive HIV-NCD chronic care models. All qualitative interviews were conducted and audio-recorded after receiving written informed consent.

Transcripts from IDIs and KIIs were coded, similar codes were categorized, and overarching themes were generated from alike categories. Findings were validated through triangulation of IDI and KII data. Qualitative data were analyzed using ATLAS.ti version 22.

Following independent analysis, quantitative and qualitative findings were compared. The Meta-inferences were systematically drawn upon knowledge, experience, and data-driven inferences derived from literature review, reflexivity of researchers, and integrated findings from the study [23].

The specific attributes of this MMR design are described with Good Reporting of A Mixed Methods Study (GRAMMS) checklist (See online supplemental material).

### Patient and Public Involvement

PLHIV were not involved in designing, reporting, or dissemination plans of this research. However, KIs were consulted during the formulation of the study regarding methodology and questionnaire designing.

### Ethics statement

The study was approved by the Institutional Ethics Committee at AIIH&PH, Kolkata (No. AIIHPH/PSM/Protocol/2020-23/012, dated: 18-01-21), and Research Oversight Committee IPGME&R, Kolkata (Memo No. IPGME&R/IEC/2021/579, dated: 27-09-2021). Permission for dissemination of the study findings taken from West Bengal State AIDS Prevention and Control Society (Memo No: HFW-28021(99)/7/2023-CST SEC-Dept. of H&FW/979, Date: 29.11.2023). All participants provided consent, assured of confidentiality and anonymity. Findings presented anonymously.

## Results

The study included 279 quantitative interviews with 98.9% response rate (279/282). Mean (± SD) age of the participants was 44.6 (± 10.5) years. More than half (62%) was male, while four participants belonged to nonbinary or other genders. The median duration of formal schooling was 7 years (IQR 0-10), and the median per capita monthly family income was Indian rupee (INR) 3333 (IQR 2000-5000). Nearly 26% of the participants were not engaged in any paid work, that included retired, unemployed, and homemakers (Table 1).

**Table 1:**
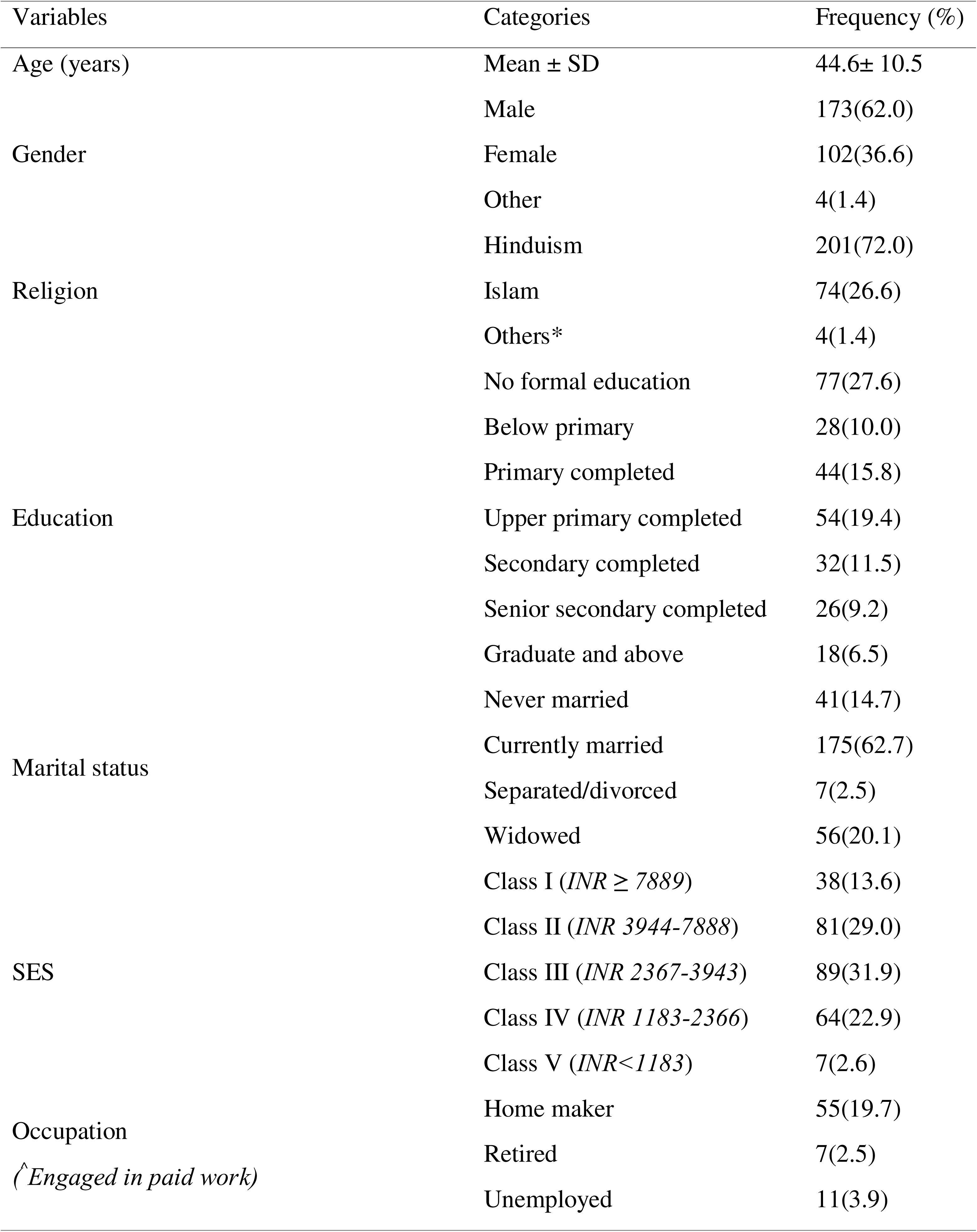
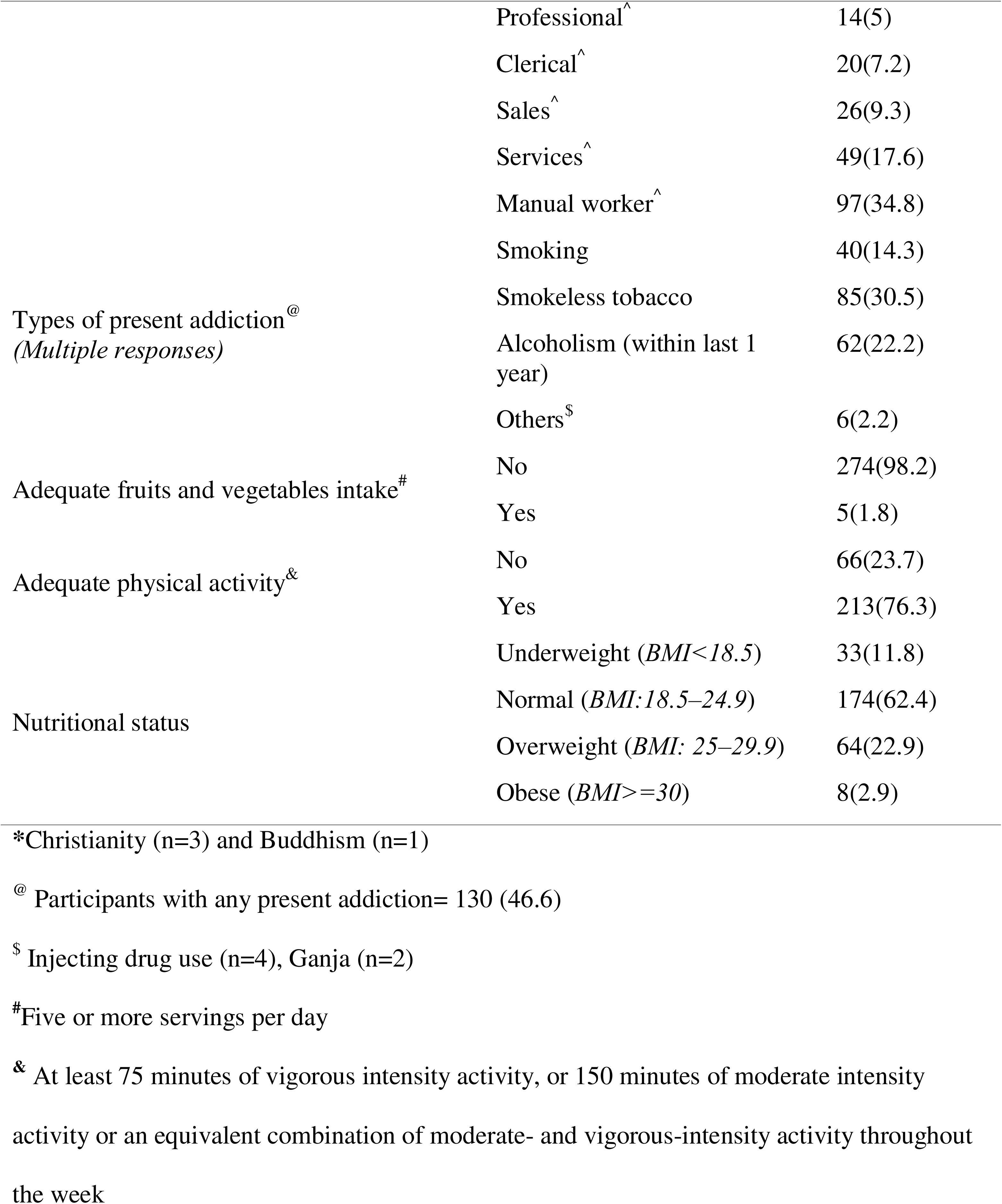
Distribution of QUAN study participants according to background characteristics and self-reported NCDs (N=279)

Almost 65% were diagnosed with HIV for more than 5 years. The medium baseline and recent CD4+ counts were 240 (152-255) and 549 (409-707) cells/mm^3^, respectively. Majority of the participants were receiving a non-Protease Inhibitor (PI)-based ART (93.9%) and had an HIV viral load <1000 copies/ml (96.0%). Over half (54.8%) of the participants reported having no health insurance coverage.

Nearly 30% (n=83) of the participants reported having at least one common NCD. DM (14.0%) was most frequently reported, followed by HTN (13.3%). Multimorbidity was present among 15 PLHIV, representing 18.1% of those having NCDs. More than half of the PLHIV suffering from NCDs (59%) sought health care services from private sector (Figure 1).

**Figure 1:**
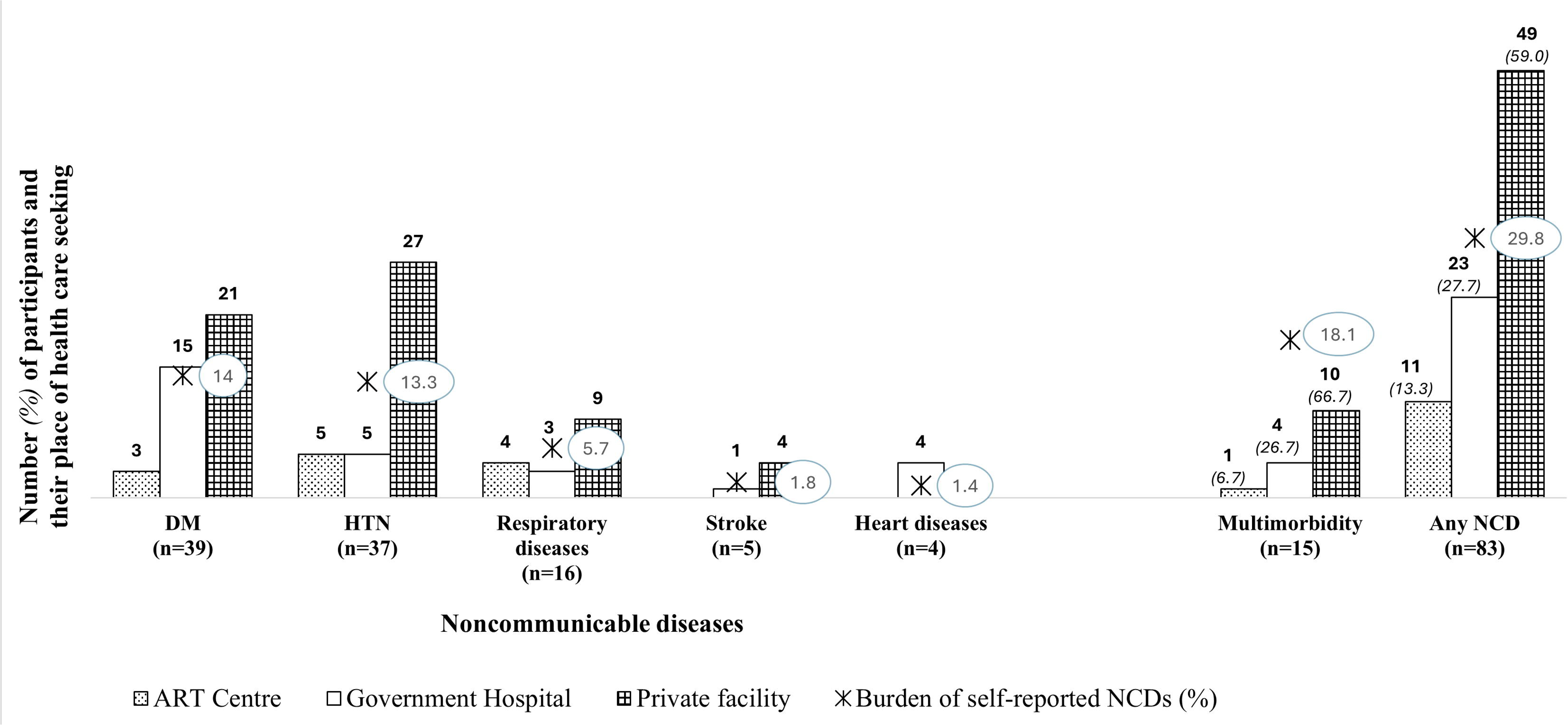
Distribution of NCDs and health care seeking behavior among the study participants (N=279)

Older age (≥45 years, aOR (95% CI) = 3.5 (1.9-6.4)), lack of adequate physical activity (2.7 (1.4-5.1)), and obesity (9.1 (1.4-58.2)) were significantly associated with higher odds of NCDs (Table 2).

**Table 2:**
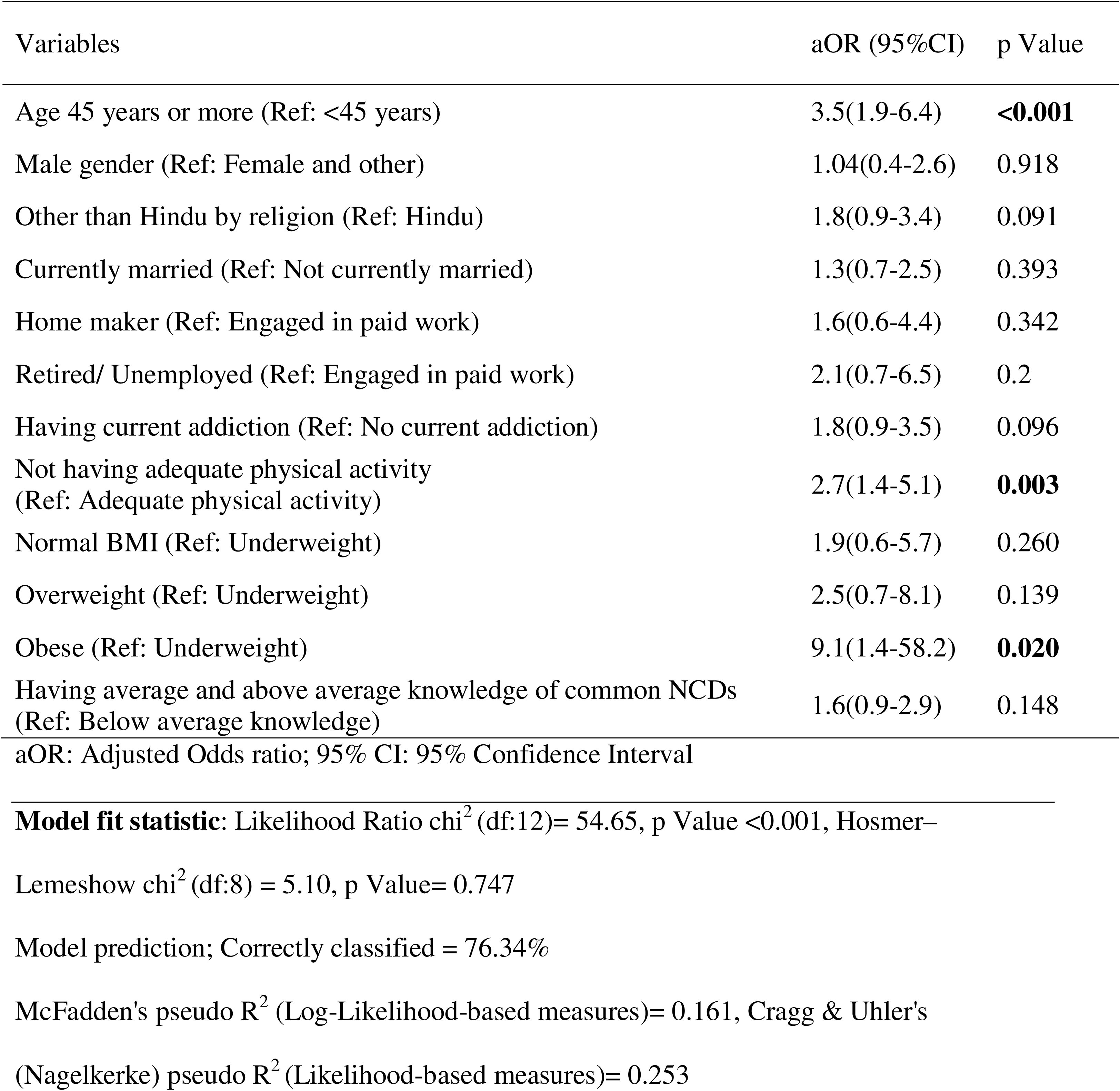
Factors associated with NCDs among the study participants: multivariable logistic regression analysis (N =279)

The qualitative strand included 10 IDI participants, aged between 30 to 65 years. The sample represented a wide educational background, from no formal schooling to qualification beyond high school education, and representative from all three genders. All were suffering from at least one NCD; seven had HTN, five had DM, and conditions like cancer, heart disease, and stroke were also documented. Half of them (5 out of 10) were living with multimorbidity and seeking treatment partially or entirely from private healthcare.

Three KIIs were conducted with a senior medical officer, a nursing officer, and a counsellor from the ART center, each possessing 5-10 years of working experience in ART service delivery. They identified DM and HTN as the most common NCDs in this population and noted that very few cancer cases have been observed among PLHIV in recent years. Codes, category, and themes were developed from the following IDIs and KIIs findings (Figure 2):

**Figure 2:**
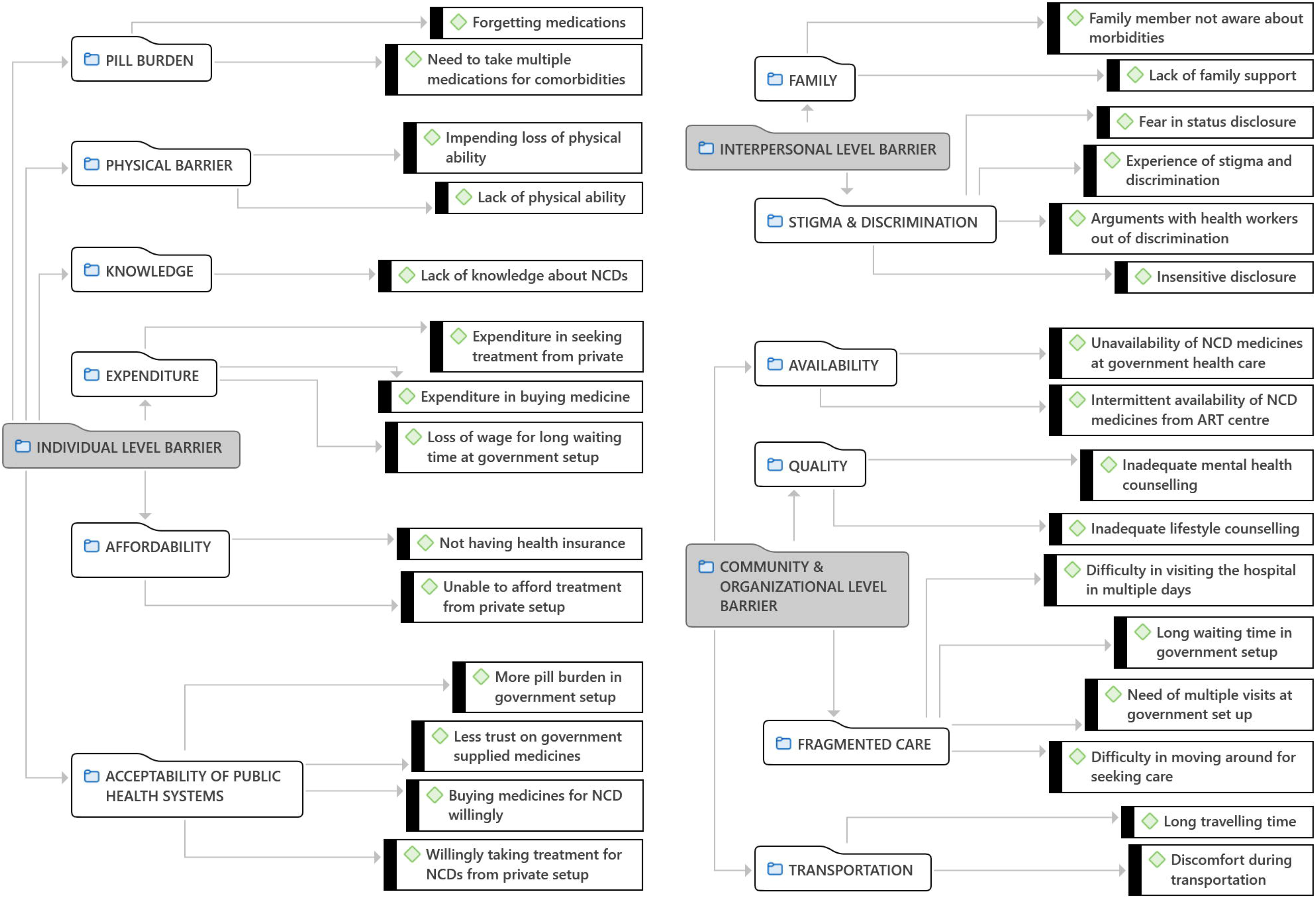
Network diagram with codes, category codes, and themes reflecting barriers to seeking health care among PLHIV with NCDs

### A. Individual Barriers-

A few notable individual level barriers included lack of physical ability, pill burden, expenditure, and issues of affordability stemming from lost wages during hospital visits, medication cost, and expense of seeking private care. Additionally, public health systems suffered from a lack of trust due to the complex, difficult-to-navigate pathways required to access care.

A participant aged between 40-50 years shared, “*I was visiting a Government Hospital, where I received numerous medications. Now that I visit a private clinic, they’ve streamlined my prescription to just three This reduction in the number of medicines has been beneficial for me”*,

Another interviewee age between 50-60 years added, “*Currently, I have to navigate different places for the treatment. This involves arranging transportation and is becoming increasingly difficult as I age, and my physical ability deteriorates*”,

One participant also mentioned, “*Going to the hospital consumes our entire day. It is time-consuming and leads to a loss of wages*”

KIs also echoed these sentiments:

> *“Individuals may experience anxiety and a perceived burden of multiple medications” “Financial constraints sometimes lead to interrupted NCD care, especially in private care” “While public facilities offer most essential medications, the process is burdensome”*

### B. Interpersonal Barriers-

Lack of family support, and fear of disclosing one’s HIV status were common, as one participant confided, *“No, I haven’t informed the doctor I see for minor issues and blood pressure checks…. I could not even tell my family members; how could I tell him”*.

HIV-related stigma and discrimination remain persistent obstacles. An interviewee aged between 30-40 years recounted, *“I experienced significant mistreatment during my hospital stay for surgery …… The staff, including nurses, attendants, and doctors, seemed reluctant to interact with me, often avoiding eye contact and conversation They were unresponsive to my questions and concerns, leaving me feeling neglected and distressed…. They used my illness as a justification for their mistreatment, implying that I deserved to be treated poorly”*.

KIs confirmed that, “*Despite considerable progress, the stigma surrounding HIV persists, with lingering concerns about casual contact and transmission a proactive approach to the care of HIV patient remains lacking……focus seems to be on short-term management rather than comprehensive, patient-centered care.”*

### C. Community and Organizational Barriers-

Systemic challenges at community and organizational level significantly impede care. Long travel times to seek healthcare, extended waiting times at government facilities, along with inconsistent medication availability, were some of the significant challenges.

An elderly participant explained, *“I am receiving treatment at another clinic within the same hospital and purchasing the medications prescribed by them. They are also recommending some further investigations, which I am doing from outside of the hospital to avoid multiple visits”,*

Another one added*, “In the past, I have received medication for diabetes and hypertension from this clinic for intermittent periods. Right now, those are not available…. The waiting times in the public healthcare system are extensive, often taking 4-5 hours just to see a doctor. The process involves queuing for tickets, separate appointments for investigations and reports, which can easily stretch a single visit across three days. For these reasons, I’m currently seeking treatment privately”*.

KIs corroborated these, *“While those with financial means can readily access private healthcare for NCDs, patients relying on public health systems face significant barriers, including long queues for appointments, tests, and collecting medication. These delays deter some patients from public systems”*

### D. Comprehensive Chronic Care for HIV-NCD

Both the PLHIV and ART service providers expressed a strong desire for comprehensive HIV-NCD care under one roof. One KI summarized, *“For PLHIV who also have NCDs, a comprehensive, integrated treatment facility offers significant advantages. By providing care under one roof, we can reduce the burden on patients, streamline their experience, and improve their adherence to treatment. This is especially crucial for the many patients in our clinic who come from lower socioeconomic backgrounds. An integrated model addresses barriers to accessing and receiving care, minimizes the need to navigate multiple departments and long queues, and reduces the potential for stigma associated with disclosing their HIV status to multiple healthcare providers.”*

The joint display of quantitative and qualitative data underscores the call for comprehensive HIV-NCD care. However, it also identifies substantial challenges to implementation (Table 3).

**Table 3:**
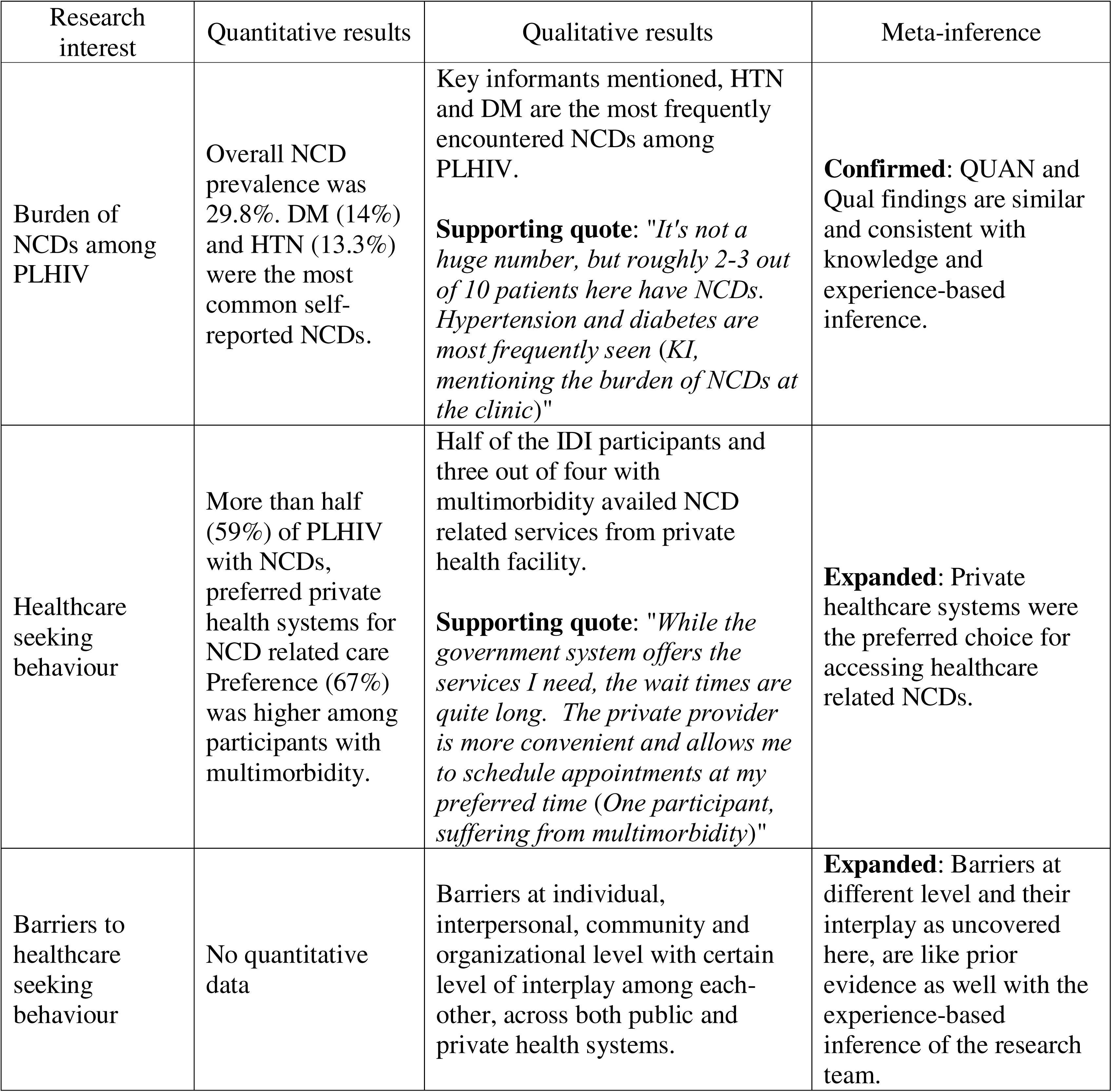
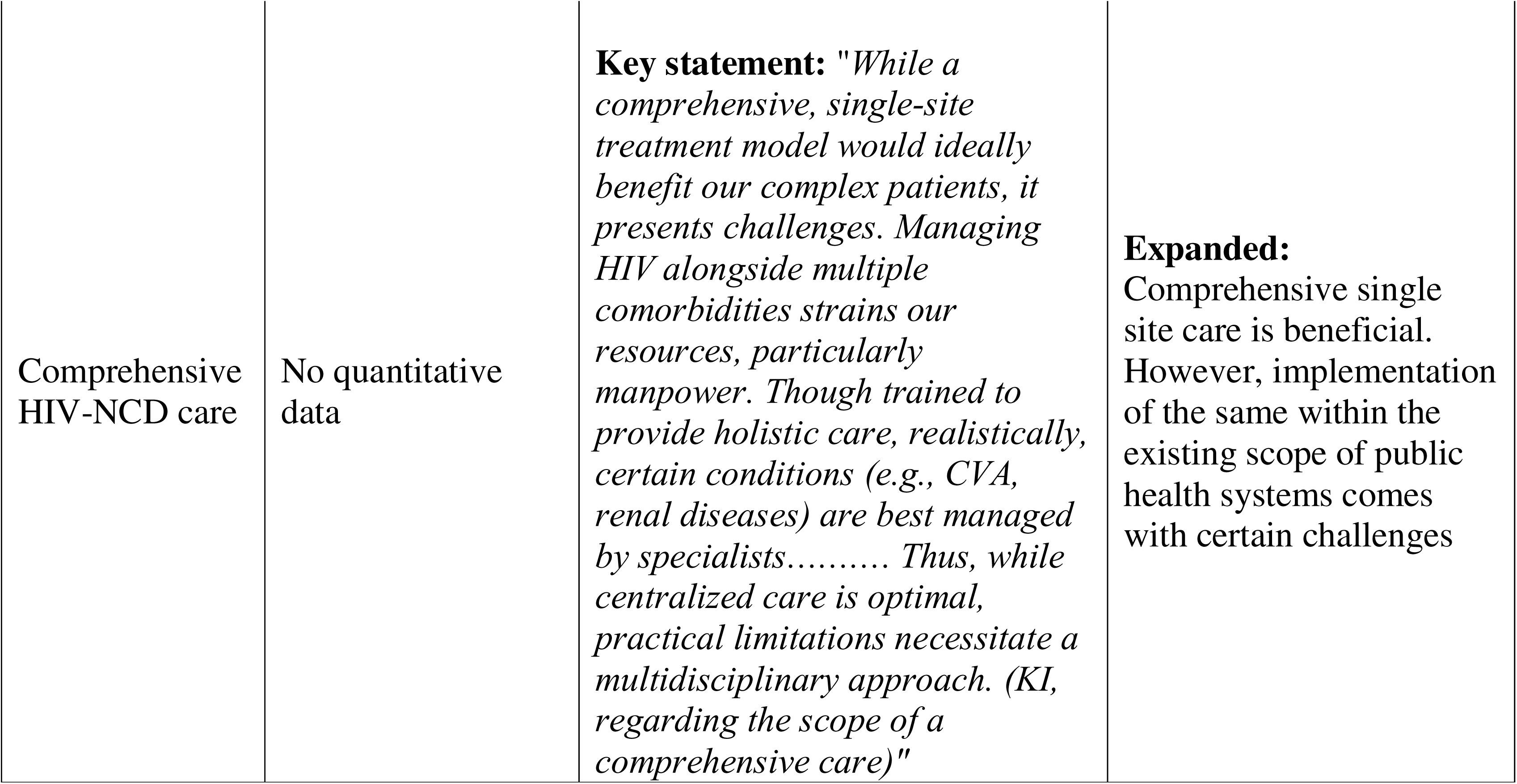
Joint display comparing the quantitative and qualitative findings side-by-side and revealing the meta inference (N_QUAN_= 279, N_IDI_ =10 & N_KII_ =3)

## Discussion

Management of NCDs among the aging population of PLHIV has become a critical aspect of HIV care. [24]. In this context the study employed a mixed-methods approach, to assess the burden and risk factors of NCDs, examine healthcare-seeking behavior, and identify barriers to care. The findings were used to discuss the feasibility of comprehensive NCD care model for PLHIV.

More than a quarter of the PLHIV were suffering from NCDs. Prevalent risk factors included tobacco chewing, smoking, alcoholism, low fruit and vegetable intake, while older age, inadequate physical activity, and obesity were identified as significant contributing factors to NCDs. Private healthcare was preferred for NCD treatment. However, PLHIV with NCDs often faced numerous barriers when accessing care, both in the public and private sectors. The findings highlighted the need for comprehensive, integrated chronic care models to address the complex health needs of PLHIV with NCDs.

The socio-demographic characteristics of the study population, particularly aging, corroborate with previous reports [12, 16, 25, 26]. Inadequate dietary habits (low fruit and vegetable consumption), lower educational attainment (fewer years of schooling), and high unemployment were consistent with the socioeconomic challenges frequently documented among PLHIV in India [25, 27].

In the absence of comparable local studies, assessing the relative NCD burden among PLHIV is challenging. Global reports indicate considerable variations in NCD burden (14-48%) due to methodological differences. This study estimate (30%) falls within the mid-range of those prior reports [3, 6, 10–12, 28–30]. However, self-reporting and the exclusion of terminally ill PLHIV, unable to attend the ART center could have led to an underestimation of the true burden [10]. This study likely represents the minimum burden of NCDs among the PLHIV, highlighting the need for more resource intense research.

A wide variation in the estimated burden of DM and HTN among PLHIV were also reported [3, 31–33]. While the present estimates fall within the reported ranges, underestimation could not be ruled out due to self-reporting, limited disease awareness, survival bias, and inadequate NCD screening. Nevertheless, DM and HTN remained the most prevalent comorbidities. The estimates for CVDs and stroke were consistent with previous reports [28, 31]. However, intensive investigations elsewhere have documented higher burdens, suggesting that targeted screening is crucial to uncovering the true magnitude [33, 34].

Paucity of the cancer cases in this study, likely reflect the limited sample size rather than the true absence of the disease, particularly given the declining prevalence of AIDS-related cancers [35].

Private healthcare systems were preferred, particularly with NCD multimorbidity. Given the lower socioeconomic status of the study participants, this trend raises concerns about fair financing – a key goal of global health systems.

Upon further exploration of the care seeking preferences, the study revealed multiple, interconnected barriers at personal, interpersonal, community, and organizational level [13–15]. Individual level barriers such as pill burden and a lack of knowledge about NCDs could be addressed through organizational measures like providing tailored counselling. Physical barriers could be aggravated by challenging public transportation and fragmented health care delivery systems compelling PLHIV to visit multiple departments over extended periods.

While strong interpersonal support from family could partially mitigate these challenges, system level interventions are ultimately needed to removing the barriers. The low acceptability of public healthcare, often resulted from fragmented services, medication shortages, and lack of health system responsiveness, compelling individuals towards private care. Such shift brings additional financial burden, issue of affordability, heightens fear of HIV status disclosure resulting in inappropriate treatment decisions by private physicians unaware of the patient’s HIV status. Unfortunately, status disclosure in public healthcare can also precipitate stigma and discrimination, further discouraging care-seeking.

While similar barriers to NCD care, such as, financial constraints, systemic insufficiencies, socio-cultural factors are observed among general population [36,37], PLHIV additionally sufferer from dual burden of chronicity of both HIV and NCD, cumulative pill burden and adverse effects of ART, HIV specific pathological changes, and certainly the distress of stigma steaming from the HIV status [38, 39].

This study has several limitations. Estimation of the burden of common NCDs, was based on the cross-sectional design relying on self-reporting, which may underestimate the burden. Further, by excluding terminally ill or bedridden patients, unable to attend the ART center, study might have missed a segment of population with higher NCD burden. The cross-sectional design also introduces survival bias and temporal ambiguity while assessing the burden and risk factors. Sample size was limited to estimation of overall NCD prevalence, but not sufficient for less common conditions, such as cancer. There was no scope to directly compare the burden of NCDs and barriers of healthcare seeking, among similar general population. Inclusion of ART service providers as KIs could introduce information bias while exploring system limitation. Finally, the researchers, being public health professionals, and having experience of working with PLHIV in similar context, could have subjective interpretation as an insider. To mitigate any assumption that could stemmed from our own experience, the research team was engaged in reflexivity. But the findings finally resonated with our own experiences, which were initially set aside by bracketing. The study was also unable to involve PLHIV in designing and reporting the research. However, within the available resources, we believe this mixed-methods approach provided an efficient exploration of the research area.

To conclude, the ageing PLHIV population carries a significant burden of NCDs. Among PLHIV with the dual burden, the chronic nature of both HIV and NCD, combined with physical inability, cumulative pill burden, expenditure, distress of stigma imposes substantial challenge to seeking NCDs care. This scenario demands implementation of comprehensive HIV-NCD care. While public health systems have the potential to offer comprehensive services to PLHIV with NCDs, fragmentation often undermines community trust. Private health care systems on the other hand offer more accessible, time efficient services for NCD care, but at the cost of out-of-pocket expenses, not affordable to many, and carry greater concern for status disclosure. Stigma and discrimination remain pervasive barriers, even affecting access to health insurance for PLHIV. This complex interplay of barriers can lead to disengagement from NCD care, resulting in increased morbidity and mortality. These data-driven conclusions strongly align with our personal observations as public health professionals, highlighting the urgent need for improved integration within the public health system. However, integrated care needs to be built upon resource availability and training of the service providers [40].

By transitioning to an integrated care model, the health system can directly resolve prominent organizational barriers of fragmented care. Consolidating ART and NCD medicine dispensation, alongside comprehensive lifestyle and mental health counselling, into single visits would minimize the physical strain, reduce long waiting times, and mitigate the loss of wages due to multiple and extended navigations in the hospitals. Furthermore, unified care delivery can reduce the risk of insensitive status disclosure by limiting the number of separate departments and healthcare workers a patient must interact.

The insights gained from this study can be used to identify systemic gaps and tailor it towards a more patient-centric approach. While NACO envisions a comprehensive chronic care model for PLHIV, implementation barriers exist. Addressing fragmentation within the existing system presents a significant opportunity to improve NCD interventions for PLHIV. Implementation research, building on these findings, is required to test new models of comprehensive HIV-NCD care utilizing available resources effectively and efficiently, within the public health systems of the country [24, 41].

## Data Availability

All data produced in the present study are available upon reasonable request to the authors

## Declaration

We hereby declare that this is our own work and there is no conflict of interest.

## Data availability

Anonymous data (after removal of direct and indirect identifier) are available on request to the corresponding author.

## Acknowledgement

Contribution of the authors to the study

Sandipta CHAKRABORTY: Conceptualization, Proposal and Tool development, data collection and entry, data analysis, manuscript-drafting, review and finalization

Shantasil PAIN: Conceptualization, Proposal and Tool development, data analysis, manuscript-review and finalization

Pankaj Kumar MANDAL: Conceptualization, Proposal and Tool development, data analysis, manuscript-review and finalization

Bobby PAUL: Conceptualization, Proposal and Tool development, data analysis, manuscript-review and finalization

Dipankar JANA: Conceptualization, Proposal and Tool development, data collection and entry, data analysis, manuscript-review and finalization

Preeti GURUNG: Conceptualization, Proposal and Tool development, data analysis, manuscript-drafting, review and finalization

The authors expressed their acknowledgement to all the PLHIV attending the ART Centre and all the healthcare service providers of the ART Centre, without their support and cooperation this study would have a distant dream.

The authors acknowledge the use of Google Gemini in language reframing of the final manuscript.

## Supplemental material

Good Reporting of A Mixed Methods Study (GRAMMS) checklist

